# Agent Role Structure and Operating Characteristics in Large Language Model Clinical Classification: A Comparative Study of Specialist and Deliberative Multi-Agent Protocols

**DOI:** 10.64898/2026.02.22.26346818

**Authors:** Callum Anderson

**Affiliations:** Faculty of Engineering, University of Ottawa, 800 King Edward Avenue, Ottawa, Ontario K1N 6N5, Canada

**Keywords:** Large language models, Multi-agent systems, Role decomposition, Structured prompting, Clinical decision support, Operating characteristics

## Abstract

Large language models (LLMs) are increasingly considered in clinical decision support, yet the architectural effects of role decomposition within multi-agent systems remain poorly isolated. Prior comparisons of single-agent and multi-agent prompting often confound workflow structure with changes in model configuration, training, or decoding. We present a controlled study evaluating whether specialist and generalist multi-agent protocols produce different operating characteristics under fixed inference conditions. Two deterministic protocols, Generic Deliberative (GD) and Feature-Specialist (FS), are evaluated under identical base weights, decoding settings, computational budget, and adjudication logic. The primary comparison isolates role structure and information routing, while feature-assignment and model-scale ablations evaluate boundary conditions. On the UCI Cleveland Heart Disease and Pima Indians Diabetes benchmarks, altering role structure produces substantial operating-characteristic shifts. Under the primary configuration (`llama3.1:8b`), Cleveland FS achieves higher accuracy (0.72 vs. 0.65) and macro-F1 (0.71 vs. 0.65), with a specificity-favoring operating point, whereas Pima GD achieves higher accuracy (0.68 vs. 0.51) and macro-F1 (0.64 vs. 0.49), while FS induces pronounced class asymmetry. Feature-pair ablations show that specialist feature assignment is a design lever, with FS accuracy ranging from 0.41 to 0.72. A model-scale ablation (`qwen2.5:14b`) shows that protocol-level effects interact with base-model capacity. Overall, internal role decomposition should not be treated as safety-neutral: it can act as a feature-conditioned architectural bias that changes evidence aggregation and shifts class-wise error profiles, indicating that prompt structure and architectural configuration should be explicitly tuned, validated, and reported in safety-sensitive LLM-based clinical decision systems.

## 1. Introduction

Recent advances in large language models (LLMs) have increased interest in their use in clinical settings. Healthcare institutions now maintain extensive structured electronic health record (EHR) repositories, and artificial intelligence techniques are increasingly being applied to these data for clinical prediction, decision support, and workflow optimization [1]. Reviews of medical natural language processing in the LLM era show that modern models encode substantial biomedical knowledge and can support clinically relevant reasoning, particularly when adapted through instruction tuning and task-specific guidance [2]. Empirical studies further demonstrate that LLMs can process structured clinical artifacts, such as eligibility criteria and patient descriptors, and produce representations that support grouping and classification tasks [3]. LLM-based systems have also been explored for identifying social and contextual patient information from clinical records and incorporating these factors into predictive workflows [4]. Together, this body of work suggests that LLMs can operate over structured clinical information and support medical classification-oriented tasks. In medical contexts, locally deployable large language models are of particular interest, as they allow sensitive patient data to remain within institutional infrastructure while maintaining operational autonomy and regulatory compliance [5]. However, despite increasing adoption of multi-agent prompting workflows, the specific effects of internal role decomposition within LLM-based decision systems remain poorly isolated. In particular, it remains unclear whether prompt-level agent specialization systematically alters classification behavior under controlled inference conditions, and how such effects depend on evidence routing and model capacity.

Efforts to improve reliability and reasoning depth in LLM-based systems have increasingly adopted multi-agent or chained architectures. Rather than relying on a single model invocation, these systems decompose complex tasks into coordinated sub-tasks executed by multiple LLM instances, often with explicit roles or sequential dependency structures [6, 7]. These approaches draw conceptual parallels to ensemble-based decision systems, where multiple decision-makers contribute complementary perspectives to reduce error and improve robustness [8]. In computational settings, structured LLM workflows have been shown to make intermediate reasoning steps explicit and to introduce validation and adjudication mechanisms between agents, thereby increasing the auditability of system behavior and allowing more deliberate control over task execution [6]. These architectural patterns are particularly relevant for safety-sensitive domains such as clinical and diagnostic decision support, where structured multi-step reasoning and cross-checking may offer advantages over single-agent architectures. In these environments, shaping operating characteristics, including tradeoffs among sensitivity, specificity, and precision, is critical, as clinical workflows often prioritize minimizing missed diagnoses even at the expense of increased false positives [9].

Although a growing number of multi-agent LLM frameworks introduce layered, cooperative, or dynamically orchestrated agent structures, most prior research has emphasized architectural design and application-level performance rather than tightly controlled analysis of internal role decomposition [6, 10, 11]. Relatively little attention has been given to isolating prompt-level role structure as an independent architectural variable under controlled experimental conditions. Therefore, it remains unclear how much internal agent specialization or overlapping full-view reasoning alone contributes to observed differences in classification behavior and operating characteristics.

To address this specific gap, we conduct a controlled comparison of two structured multi-agent architectures that differ in internal role decomposition and patient data access. Both are implemented as deterministic Directed Acyclic Graph (DAG) workflows operating on two tabular clinical benchmarks, the UCI Cleveland heart disease dataset [12] and the Pima Indians Diabetes dataset [13], with agents running in parallel prior to a downstream adjudication step. In the GD configuration, generalist agents independently evaluate the complete feature set, whereas in the FS configuration, agents are assigned distinct clinically relevant feature domains and produce complementary partial assessments. Intermediate evidence representations are standardized across architectures, and model weights, decoding temperature, computational budget, aggregation logic, and implementation code are held constant in the primary comparison. This design isolates role structure and information routing, enabling direct assessment of how internal agent organization shifts classification performance and redistributes sensitivity–specificity trade-offs across distinct clinical prediction tasks. We further test the robustness and boundary conditions of these effects through feature-assignment and model-scale ablations, evaluating the extent to which FS behavior is influenced by selected specialist features and whether GD–FS differences persist under a larger base model.

## 2. Methods

### 2.1. Datasets

We evaluate both architectures on two structured clinical benchmarks originally released through the UCI Machine Learning Repository: the Cleveland Heart Disease dataset [12] and the Pima Indians Diabetes dataset [13]. Both datasets are widely used for binary disease-status classification and contain tabular patient-level clinical measurements.

The processed Cleveland subset contains 303 patient records with 13 structured clinical features spanning demographic variables, exercise test results, and imaging-derived indicators. The positive class prevalence in the binarized Cleveland dataset is 0.459 (45.9%).

The Pima Indians Diabetes dataset contains 768 patient records with 8 structured clinical features including laboratory measurements, anthropometric indicators, and family history scores. The positive class prevalence in the Pima dataset is 0.349 (34.9%).

For consistency across experiments, both datasets use a unified binary target variable `disease_present`, where 0 denotes absence of disease and 1 denotes disease presence. Although the semantic interpretation differs by dataset, representing cardiovascular disease in Cleveland and diabetes in Pima, the encoding is identical, enabling controlled comparison of architectural effects across tasks.

Throughout this study, model outputs are interpreted as binary disease-status classification labels for benchmark datasets, not as clinical diagnoses or patient-level medical decisions.

#### 2.1.1. Preprocessing and Labeling

For the Cleveland dataset, the original target variable (`num`), which encodes heart disease severity on an ordinal scale from 0 to 4, was binarized. Records with a value of 0 were mapped to `disease_present` value 0 (no disease), whereas records with values 1 through 4 were mapped to value 1 (disease present).

The Pima dataset natively provides a binary outcome variable (`Outcome`), which was directly mapped to the unified `disease_present` target without modification.

For the Cleveland dataset, missing values were handled via per-feature mode imputation computed across the processed subset. For each variable containing missing observations, the most frequent observed value was used to replace null entries. Mode imputation was selected because the missing Cleveland fields were encoded categorical variables, for which mean imputation is not semantically meaningful, and because the small benchmark setting favored a deterministic preprocessing rule over an additional model-based imputation procedure such as KNN. However, mode imputation may bias imputed records toward the majority category, and an LLM could amplify this bias when interpreting clinically salient imputed features.

The Pima dataset contains zero-coded entries in certain physiological variables that are commonly treated as missing in distributed versions of the dataset. In this study, all features were preserved in their original encoded form and no additional missing-value indicators were introduced into the serialized prompts presented to the language model. This choice preserved the public benchmark representation and avoided introducing an additional preprocessing intervention across features. However, physiologically implausible zero values may introduce noise into the serialized patient records and could be interpreted literally by the LLM.

Each dataset was serialized into a structured textual representation for LLM input. Cleveland features include `age_years`, `sex`, `chest_pain_type`, `resting_blood_ pressure_mmHg`, `serum_cholesterol_mg_dL`, `fasting_ blood_sugar_over_120_mg_dL`, `resting_ecg_result`, `max _ heart _ rate _ bpm`, `exercise _ induced _ angina`, `st _ depression _ exercise _ relative _ to _ rest`, `st _ segment_slope_peak_exercise`, `num_major_vessels_ fluoroscopy`, and `thalassemia_test_result`.

For the Pima Indians Diabetes dataset, no feature transformations were applied beyond renaming columns to standardized LLM-facing identifiers while preserving all original numeric values. Pima features include `pregnancies`, `plasma_glucose_mg_dL`, `diastolic_ blood_pressure_mmHg`, `triceps_skinfold_thickness_ mm`, `serum_insulin_uU_mL`, `body_mass_index`, `diabetes_ pedigree_function`, and `age_years`. A complete mapping of raw variables, encoded values, and LLM-facing feature names for both datasets is provided in Appendix B.

### 2.2. Prompting Protocols

We implement two structured multi-agent protocols that share identical model configuration, decoding parameters, adjudication logic, and a common structured evidence representation consisting of signal, strength, and justification fields. Both protocols operate as DAG workflows in which upstream agents generate structured intermediate assessments that are consumed by a downstream adjudicator. The protocols differ only in internal role decomposition and information routing. The terms “clinician,” “doctor,” “specialist,” and “adjudicator” are used as prompt-role labels to structure evidence review and aggregation; they do not denote real clinical professional roles, clinical accountability, or a deployable diagnostic workflow.

In addition to the two multi-agent protocols, we evaluate a single-agent baseline as a non-decomposed reference point. In this baseline, a single agent receives the full patient record and produces the final disease-status classification directly in one LLM call, under the same base model, deterministic decoding settings, and output format as the multi-agent protocols. The single-agent baseline is included only as a reference for the multi-agent comparisons and is not subjected to the feature-assignment or model-scale ablations.

#### 2.2.1. Protocol A: Feature-Specialist Architecture (FS)

The Feature-Specialist (FS) architecture comprises two parallel specialist agents, each restricted to a single clinically defined feature from the patient record. Each specialist produces a structured opinion describing the evidentiary contribution of its assigned feature, including (1) a directional signal (`supports_disease`, `supports_no_disease`, or `unclear`), (2) a categorical strength rating (`low`, `medium`, `high`), and (3) a brief justification note explicitly referencing the feature value. Specialists operate independently and are prohibited from reasoning over features outside their assigned scope.

The primary FS feature pairs were selected to provide two non-overlapping, interpretable evidence channels for each dataset, rather than to identify an optimal feature subset. For Cleveland, specialists were assigned `chest_ pain_type` and `num_major_vessels_fluoroscopy`. These correspond to a symptom-presentation feature and a test-derived anatomical feature, respectively; both are part of the standard Cleveland schema and have been retained among selected predictors in prior heart-disease prediction work using the Cleveland dataset [14]. For Pima Indians Diabetes, specialists were assigned `plasma_glucose_mg_dL` and `body_mass_index`, pairing a direct glycaemic measurement with a body-measurement risk factor. Prior Pima feature-selection and prediction analyses identify glucose and BMI as important predictors for diabetes classification [15]. Thus, the initial FS configuration routes two distinct and individually informative signals through separate specialist roles while preserving interpretability. Sensitivity to these initial assignments is evaluated through the feature-assignment ablation in Section 3.4.

Formally, given a serialized patient record *x*, each specialist receives the record but is instructed to evaluate only its assigned feature *ϕ*_*i*_. Each specialist then produces an opinion

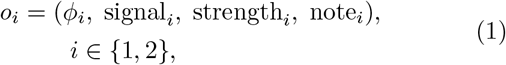

where *ϕ*_*i*_ denotes the feature assigned to specialist *i*. The adjudicator then receives the full record and both opinions and outputs the final binary prediction:

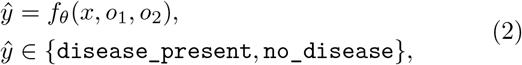

where *f*_*θ*_ denotes a third LLM call under fixed model weights and decoding settings. The adjudicator is instructed to treat strength as ordinal evidence (high *>* medium *>* low); when specialist evidence conflicts or is weak or unclear, it may consult the full record to decide. If genuinely uncertain, `no_disease` is used as a conservative tie-break. If any agent fails to produce valid JSON, a conservative fallback output is substituted. All instances of fallback utilization are transparently reported in the Results section and fully logged in the public Git repository accompanying this study.

As shown in Figure 1, the FS protocol assigns each upstream agent a single feature and routes their structured outputs to an adjudicator.

**Figure 1:**
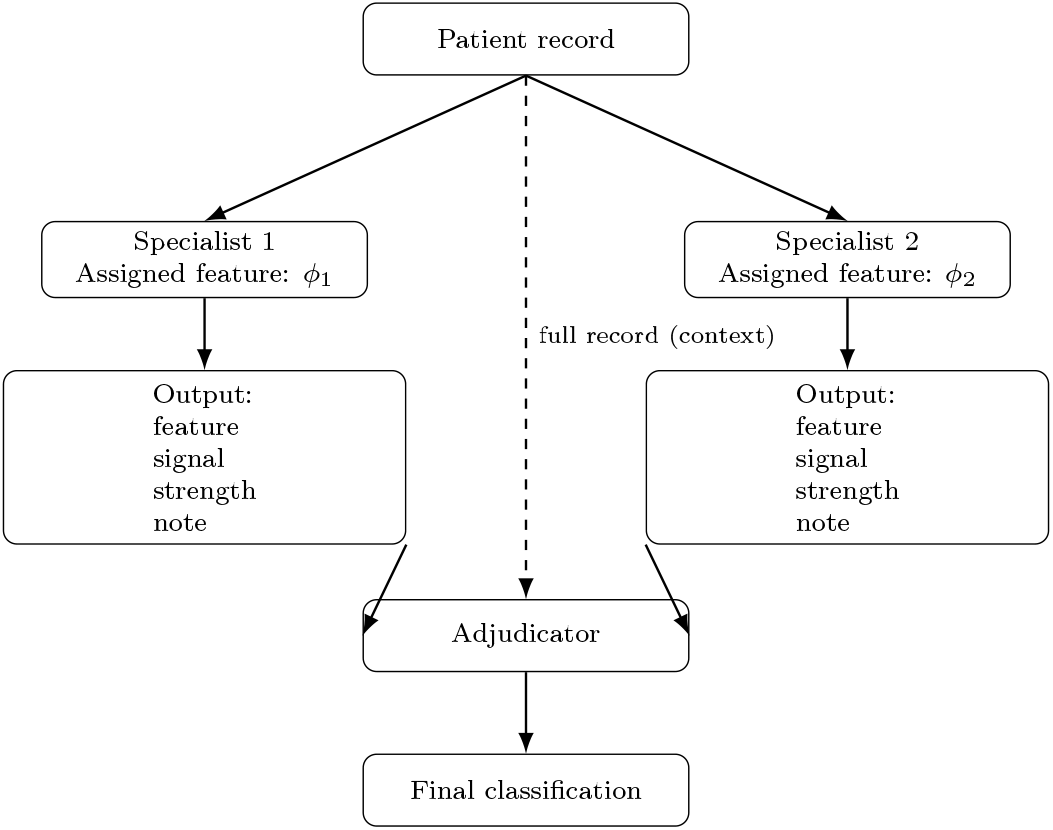
Feature-specialist (FS) architecture. Two agents independently evaluate distinct assigned features (*ϕ*_1_, *ϕ*_2_) from the patient record. Their structured outputs (feature, signal, strength, note) are routed to an adjudicator, which also receives the full record and produces the final disease-status classification. The specific feature assignments are dataset-dependent.

#### 2.2.2. Protocol B: Generic Deliberative Architecture (GD)

The Generic Deliberative (GD) architecture comprises two parallel generalist clinician agents, each evaluating the full patient record. Each clinician produces a structured opinion consisting of (1) a directional signal (`supports_disease`, `supports_no_disease`, or `unclear`), (2) a categorical strength rating (`low`, `medium`, `high`), and (3) a brief justification citing one to two key record-level factors. The clinicians operate independently without inter-agent communication.

Formally, given a serialized patient record *x*, each clinician produces an opinion

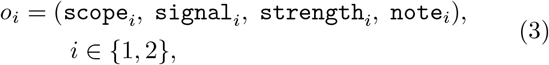

where `scope`_*i*_ = `overall_record` for both clinicians, indicating that each agent evaluates the complete patient record rather than a restricted feature subset. The adjudicator then receives the full record and both opinions and outputs the final binary prediction:

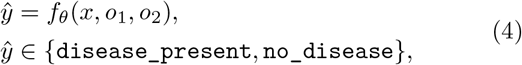

where *f*_*θ*_ denotes a third LLM call under fixed model weights and decoding settings. The adjudicator is instructed to treat strength as ordinal evidence (high *>* medium *>* low); when clinician evidence conflicts or is weak or unclear, it may consult the full record to decide. If genuinely uncertain, `no_disease` is used as a conservative tie-break, and the adjudicator is explicitly instructed not to systematically favor either clinician. If any agent fails to produce valid JSON, a conservative fallback output is substituted. All instances of fallback utilization are transparently reported in the Results section and fully logged in the public Git repository accompanying this study.

Figure 2 shows the GD protocol, in which both upstream agents evaluate the full record and provide independent opinions to the adjudicator.

**Figure 2:**
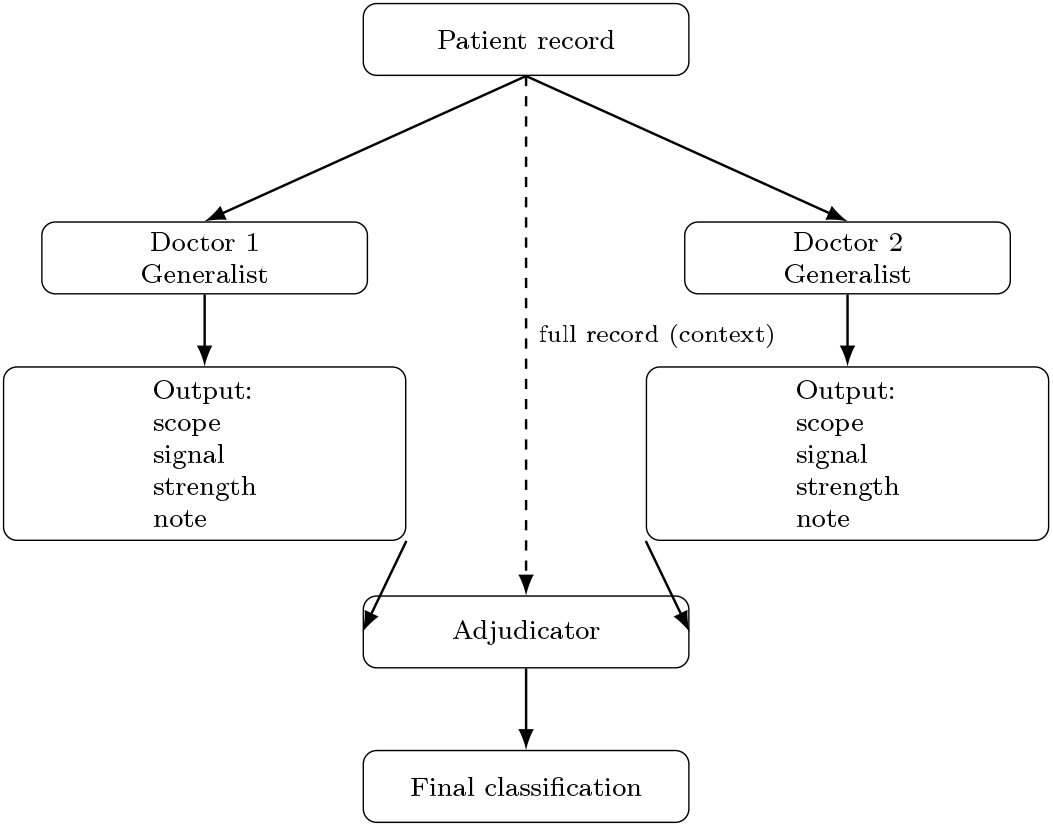
Generic deliberative (GD) architecture. Two generalist clinician agents independently evaluate the full patient record and produce structured outputs (scope, signal, strength, note), where `scope` is set to `overall_record`. These outputs are routed to an adjudicator, which also receives the full record and produces the final disease-status classification.

### 2.3. Ablation Design

To assess whether the observed differences between the Generic Deliberative (GD) and Feature-Specialist (FS) protocols were robust to design choices, we conducted two ablation analyses. The first evaluated sensitivity to specialist feature assignment within the FS protocol. The second evaluated sensitivity to base-model scale using the original GD and FS configurations.

#### 2.3.1. Feature-Assignment Sensitivity Analysis

To evaluate whether FS behavior depended on the primary specialist feature pair, we repeated the FS protocol using alternative feature assignments for each dataset. Only the two specialist feature assignments were varied; the FS architecture, prompts, adjudication logic, base model, decoding settings, fallback behavior, and evaluation procedure were held fixed.

The original FS pair served as condition A. Three additional conditions were defined to span distinct evidence-routing regimes: B, a high-signal non-overlapping alternative pair; C, a weak-signal pair intended to test the lower bound of FS behavior; and D, a mixed-signal pair combining a stronger predictor with a weaker or noisier co-specialist feature.

For Cleveland, the alternative pairs were `thalassemia_ test_result` with `st_depression_exercise_relative_ to_rest` (B), `fasting_blood_sugar_over_ 120 _mg_ dL` with `serum _ cholesterol_ mg _ dL` (C), and `max _ heart_rate_bpm` with `thalassemia_test_result` (D). For Pima Indians Diabetes, the alternative pairs were `age_years` with `diabetes_pedigree_function` (B), `diastolic_blood_pressure_mmHg` with `triceps_ skinfold_thickness_mm` (C), and `plasma_glucose_mg_ dL` with `serum_insulin_uU_mL` (D).

#### 2.3.2. Model-Scale Sensitivity Analysis

To evaluate whether protocol-level effects were specific to the original base model, we conducted a limited model-scale sensitivity analysis by repeating the original GD and FS configurations using `qwen2.5:14b`. The prompts, feature assignments, adjudication logic, decoding settings, fallback behavior, and evaluation procedure were preserved; only the base model was changed. This experiment was intended to test whether GD–FS differences persisted under a larger open-weight model, rather than to provide a comprehensive scaling analysis across model families or parameter ranges.

### 2.4. Implementation and Reproducibility

All experiments were implemented in Python 3.13, with multi-agent orchestration handled via LangChain and local inference performed through Ollama. Some ablation experiments were executed on rented GPU infrastructure via RunPod to reduce wall-clock runtime. The computational environment did not alter the experimental protocol: model identifiers, prompts, deterministic decoding settings, feature assignments, adjudication logic, fallback behavior, and evaluation scripts were held fixed across local and cloud-executed runs. In the primary GD–FS comparison, all architectures used identical base model weights and deterministic decoding settings to isolate internal role structure as the manipulated architectural variable. In the feature-assignment ablations, only the FS specialist feature assignments were varied. In the model-scale ablation, only the base model was changed. The primary experiments used `llama3.1:8b`, executed locally with temperature set to zero; the model-scale ablation additionally evaluated `qwen2.5:14b`.

All source code, prompt templates, and evaluation scripts are publicly available in the associated GitHub repository:

https://github.com/CallumGA/llm-multi-agent-role-decomposition

The datasets used in this study are publicly available:

- UCI Heart Disease dataset (Cleveland subset): https://archive.ics.uci.edu/dataset/45/heart+disease
- Pima Indians Diabetes dataset (UCI-origin, accessed via Kaggle): https://www.kaggle.com/datasets/uciml/pima-indians-diabetes-database

### 2.5. Evaluation

For all evaluated systems, we report standard classification metrics, including overall `Accuracy` and macro-averaged `Precision`, `Recall`, and `F1-score`. For the primary protocol comparison, we also report class-wise metrics for the single-agent baseline, GD, and FS architectures, and confusion matrices for the GD and FS architectures, to provide a transparent breakdown of error distributions. For the ablation analyses, we report compact summaries focused on accuracy, macro-F1, sensitivity, and specificity, which capture the main operating-characteristic shifts. To characterize operating-point behavior, sensitivity is defined as recall for the `disease_present` class, and specificity is defined as recall for the `no_disease` class.

Because each deterministic protocol was evaluated once over the full available benchmark dataset under fixed decoding settings, metrics are reported as observed full-dataset performance values rather than resampling-based estimates with confidence intervals.

## 3. Results

### 3.1. Overall Performance

Table 1 summarizes overall macro performance across the three protocols.

**Table 1:**
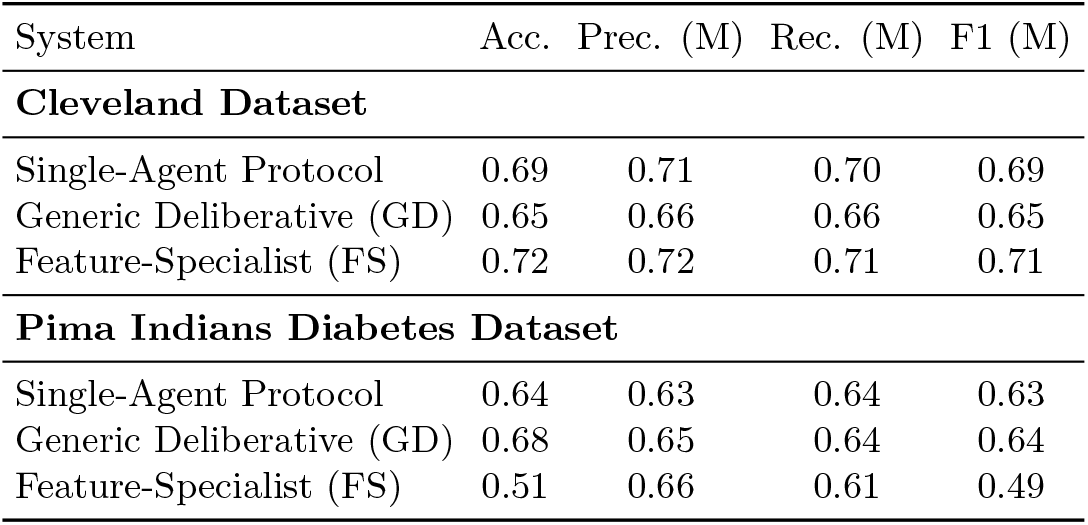
Overall performance across evaluation datasets.

| System | Acc. | Prec. (M) | Rec. (M) | F1 (M) |
| --- | --- | --- | --- | --- |
| **Cleveland Dataset** |  |  |  |  |
| Single-Agent Protocol | 0.69 | 0.71 | 0.70 | 0.69 |
| Generic Deliberative (GD) | 0.65 | 0.66 | 0.66 | 0.65 |
| Feature-Specialist (FS) | 0.72 | 0.72 | 0.71 | 0.71 |
| **Pima Indians Diabetes Dataset** |  |  |  |  |
| Single-Agent Protocol | 0.64 | 0.63 | 0.64 | 0.63 |
| Generic Deliberative (GD) | 0.68 | 0.65 | 0.64 | 0.64 |
| Feature-Specialist (FS) | 0.51 | 0.66 | 0.61 | 0.49 |

### 3.2. Class-wise Performance

Table 2 reports class-wise precision, recall, and F1-score across datasets and protocols.

**Table 2:** Class-wise performance across evaluation datasets.

| System | Class | Prec. | Rec. | F1 |
| --- | --- | --- | --- | --- |
| **Cleveland Dataset** |  |  |  |  |
| Single-Agent | no_disease | 0.80 | 0.57 | 0.66 |
|  | disease_present | 0.62 | 0.83 | 0.71 |
| Generic Deliberative (GD) | no_disease | 0.72 | 0.60 | 0.65 |
|  | disease_present | 0.60 | 0.72 | 0.66 |
| Feature-Specialist (FS) | no_disease | 0.70 | 0.82 | 0.76 |
|  | disease_present | 0.74 | 0.59 | 0.66 |
| **Pima Indians Diabetes Dataset** |  |  |  |  |
| Single-Agent | no_disease | 0.76 | 0.66 | 0.71 |
|  | disease_present | 0.49 | 0.61 | 0.54 |
| Generic Deliberative (GD) | no_disease | 0.74 | 0.79 | 0.77 |
|  | disease_present | 0.55 | 0.48 | 0.51 |
| Feature-Specialist (FS) | no_disease | 0.91 | 0.27 | 0.42 |
|  | disease_present | 0.41 | 0.95 | 0.57 |

### 3.3. Confusion Matrices

Table 3 reports confusion matrix counts (rows = true class, columns = predicted class) for the multi-agent protocols across datasets.

**Table 3:** Confusion matrices (rows = true class, columns = predicted class) across evaluation datasets.

| System | TN | FP | FN | TP |
| --- | --- | --- | --- | --- |
| **Cleveland Dataset** |  |  |  |  |
| Generic Deliberative (GD) | 98 | 66 | 39 | 100 |
| Feature-Specialist (FS) | 135 | 29 | 57 | 82 |
| **Pima Indians Diabetes Dataset** |  |  |  |  |
| Generic Deliberative (GD) | 396 | 104 | 139 | 129 |
| Feature-Specialist (FS) | 135 | 365 | 14 | 254 |

### 3.4. Feature-Assignment Sensitivity Analysis

To test whether FS behavior depended on the original specialist feature pair, we repeated the FS protocol using alternative feature assignments while holding all other protocol components fixed. As summarized in Table 4, FS performance varied substantially across the retained feature-pair conditions. The high-signal, weak/noisy, and mixed-signal assignments produced distinct sensitivity–specificity profiles, indicating that FS does not impose a fixed operating point. Instead, FS behaves as a feature-conditioned architecture whose behavior depends on the evidence channels assigned to specialists.

**Table 4:** Feature-Specialist feature-assignment ablation summary. Specificity is recall for no_disease; sensitivity is recall for disease_present.

| Dataset | Condition | Acc. | Macro-F1 | Spec. | Sens. |
| --- | --- | --- | --- | --- | --- |
| Cleveland | A — Original FS pair | 0.72 | 0.71 | 0.82 | 0.59 |
| Cleveland | B — High-signal alt. | 0.62 | 0.62 | 0.47 | 0.80 |
| Cleveland | C — Weak/noisy pair | 0.45 | 0.33 | 0.02 | 0.96 |
| Cleveland | D — Mixed-signal pair | 0.41 | 0.40 | 0.29 | 0.55 |
| Pima | A — Original FS pair | 0.51 | 0.49 | 0.27 | 0.95 |
| Pima | B — High-signal alt. | 0.45 | 0.43 | 0.19 | 0.75 |
| Pima | C — Weak/noisy pair | 0.42 | 0.41 | 0.28 | 0.67 |
| Pima | D — Mixed-signal pair | 0.48 | 0.47 | 0.48 | 0.49 |

### 3.5. Model-Scale Sensitivity Analysis

To assess whether the observed protocol-level effects were specific to the original 8B base model, we repeated the original GD and FS configurations using `qwen2.5:14b`. All prompts, feature assignments, adjudication logic, decoding settings, fallback behavior, and evaluation procedures were preserved; only the base model was changed.

As summarized in Table 5, protocol-level effects varied with base-model scale. This indicates that GD–FS differences should be interpreted as depending jointly on role structure and model capacity.

**Table 5:** Main-paper summary of model-scale sensitivity for original GD and FS configurations. Specificity is recall for no_disease; sensitivity is recall for disease_present.

| Dataset | System | Acc. | Macro-F1 | Spec. | Sens. |
| --- | --- | --- | --- | --- | --- |
| Cleveland | GD, 8B | 0.65 | 0.65 | 0.60 | 0.72 |
| Cleveland | FS, 8B | 0.72 | 0.71 | 0.82 | 0.59 |
| Cleveland | GD, 14B | 0.72 | 0.70 | 0.89 | 0.52 |
| Cleveland | FS, 14B | 0.56 | 0.46 | 0.92 | 0.14 |
| Pima | GD, 8B | 0.68 | 0.64 | 0.79 | 0.48 |
| Pima | FS, 8B | 0.51 | 0.49 | 0.27 | 0.95 |
| Pima | GD, 14B | 0.73 | 0.67 | 0.89 | 0.44 |
| Pima | FS, 14B | 0.69 | 0.69 | 0.63 | 0.81 |

Across all saved prediction outputs, no JSON fallback cases occurred: `fallback_used=True` was not observed in any primary, feature-assignment ablation, or model-scale prediction file for either dataset. The fallback mechanism was retained as a last-resort safety net for invalid or unparsable model outputs.

## 4. Discussion

### 4.1. Interpretation of Principal Findings

The performance differences between the Feature-Specialist (FS) and Generic Deliberative (GD) protocols indicate that internal role structure influences how clinical evidence is combined during inference. On the Cleveland dataset, FS achieves higher overall accuracy (0.72 vs. 0.65) and macro-F1 (0.71 vs. 0.65), but these improvements reflect a redistribution of errors rather than uniform gains. As shown in Table 3, FS substantially increases true negatives (135 vs. 98) and reduces false positives (29 vs. 66), indicating improved identification of patients without disease. This pattern is accompanied by fewer true positives (82 vs. 100) and more false negatives (57 vs. 39), reflecting lower sensitivity and a shift toward greater specificity. One explanation is that separating feature evaluation constrains how positive evidence accumulates: when specialists reason over restricted inputs, weak or ambiguous signals may be less likely to combine into an overall positive classification. In contrast, the GD protocol evaluates features jointly within a single reasoning trajectory, allowing multiple modest indicators to interact and collectively influence the prediction.

On the Pima Indians Diabetes dataset, structural effects again emerge, but manifest differently. Here, GD achieves the strongest overall macro performance (accuracy 0.68, macro-F1 0.64), whereas FS produces highly asymmetric class behavior. The FS protocol yields very high recall for the positive class (0.95) but very low recall for the negative class (0.27), corresponding to a large increase in false positives (365 vs. 104) relative to GD. Thus, in this setting, role decomposition shifts the operating point toward high sensitivity and reduced specificity, the opposite direction observed on Cleveland. The Pima dataset consists primarily of continuous numerical features, which may influence how evidence is partitioned and aggregated across agents. However, the central finding concerns within-dataset comparisons: in both benchmarks, holding model weights, decoding settings, and computational budget fixed, altering only internal role structure produces systematic changes in sensitivity–specificity trade-offs.

While it is expected that prompt variations can influence LLM outputs, the magnitude and systematic nature of the operating-point shifts observed here under deterministic conditions indicate that internal role decomposition can materially alter classification behavior. In the primary GD–FS comparison, these differences arise despite identical base model weights, decoding settings, computational budget, and adjudication logic, supporting the interpretation that role organization can function as a structured architectural bias rather than merely a superficial prompting variation. The ablation results further show that this bias is conditional rather than fixed: its direction and magnitude depend on specialist feature assignment and base-model capacity.

In clinical contexts, such trade-offs are operational rather than purely statistical. Reductions in false positives may decrease unnecessary follow-up testing or patient anxiety, whereas increases in false negatives may delay diagnosis or treatment initiation. The appropriate balance depends on intended use, such as screening versus confirmatory decision support. Overall, these findings demonstrate that multi-agent role structure does not simply improve or degrade performance globally; instead, it systematically reshapes the error profile. Careful evaluation of this redistribution is therefore essential when deploying structured LLM systems in safety-sensitive decision environments [16].

For example, a specificity-favoring configuration such as the Cleveland FS profile may be preferable in downstream confirmatory or resource-constrained triage settings where false positives carry meaningful operational costs, such as unnecessary specialist referral, additional imaging, invasive testing, or patient anxiety after an initial screening stage. In contrast, a sensitivity-favoring configuration such as the Pima FS profile may be preferable in early screening or safety-net triage, where the priority is to avoid missed cases and route potentially at-risk patients for follow-up testing, even at the expense of additional false positives. A GD-style architecture may be more appropriate when the task requires integration of multiple weak or interacting signals across the full record, whereas an FS-style architecture may be useful when specific evidence channels are trusted, clinically interpretable, and intentionally emphasized. These examples are illustrative rather than deployment recommendations, but they demonstrate why architecture-induced sensitivity–specificity shifts may be operationally meaningful in clinical decision-support design.

### 4.2. Interpretation of Ablation Findings

The feature-assignment ablations directly address whether the Feature-Specialist (FS) results are attributable only to the originally selected specialist features. The results show that FS behavior is highly sensitive to the evidence channels assigned to the specialist agents. Across both datasets, changing only the specialist feature pair substantially altered accuracy, macro-F1, specificity, and sensitivity while preserving the same architecture, prompts, adjudication logic, decoding settings, and base model. This indicates that FS should not be interpreted as imposing a single fixed operating point. Rather, FS acts as a feature-conditioned role-decomposition architecture: its operating characteristics depend on what evidence is routed to the specialists.

This finding qualifies the architectural interpretation of FS. The FS protocol does not provide a universally superior alternative to GD, nor does it produce a stable directional shift across all feature assignments. Instead, the effect introduced by FS is conditional on the constrained information channels available to the specialist agents. In Cleveland, the original FS pair produced a specificity-favoring profile, whereas the high-signal alternative shifted the system toward higher sensitivity. Weak or noisy feature pairs degraded performance and, in some cases, produced near-degenerate class bias. A similar pattern was observed in Pima, where FS favored sensitivity under the original and weak/noisy assignments, while the mixed-signal pair produced a more balanced sensitivity–specificity profile. These results suggest that role structure matters, but its effect is mediated by the quality, complementarity, and task relevance of the specialist features.

The model-scale ablation further shows that protocol-level effects interact with base-model capacity. When the original GD and FS configurations were repeated with `qwen2.5:14b`, the relative behavior of the protocols changed. On Cleveland, the larger model improved GD performance but reduced FS performance relative to the 8B configuration, producing a stronger specificity-favoring FS profile with markedly reduced sensitivity. This apparent FS collapse is therefore best interpreted as an over-conservative operating shift rather than random failure: under the restricted original Cleveland feature pair, the 14B model preserved very high specificity but rarely accumulated sufficient positive evidence to predict `disease_present`, causing sensitivity to fall sharply. On Pima, both GD and FS improved at 14B, and FS became substantially more balanced than under the 8B model while retaining higher sensitivity than GD. These results indicate that the observed GD–FS differences are not solely properties of the prompt architecture in isolation, but emerge from an interaction among role structure, feature assignment, adjudication design, and model capacity.

Although the present model-scale analysis does not include 70B+ or frontier-scale models, the 14B results suggest that role-structure effects are unlikely to disappear simply with increased capacity. Rather, we hypothesize that larger models may preserve the qualitative sensitivity of multi-agent systems to role structure and evidence routing, while reducing some failures caused by weak instruction following or poor integration of partial evidence. Under this interpretation, frontier models might diminish extreme or near-degenerate behavior in some FS configurations, but would not necessarily eliminate architecture-induced operating-point shifts. In fact, stronger models may make such shifts more controllable: if they better respect specialist-role boundaries and adjudication instructions, then the chosen evidence-routing scheme could more reliably shape sensitivity–specificity trade-offs. This remains an empirical question, and broader evaluation across larger models and model families is required before determining whether role-decomposition effects persist, amplify, or diminish at frontier scale.

Overall, the ablation results show that role-structure effects are substantial but conditional. Internal role decomposition can reshape class-wise error profiles under fixed inference conditions, but the direction and magnitude of this effect depend on specialist feature assignment and base-model capacity. This supports interpreting role decomposition and information routing as configurable architectural design choices rather than incidental implementation details.

### 4.3. Translation to Real-World EHR Settings

Although the Cleveland and Pima benchmarks provide controlled tabular settings for isolating architectural effects, real-world EHR data are substantially more complex. Contemporary EHRs are heterogeneous, large-scale data sources that combine structured clinical information with unstructured free-text documentation, including clinical notes and discharge summaries [17]. In this setting, feature-specialist design would require even more careful engineering than in the present benchmark experiments. A specialist architecture would need to weigh feature inclusion carefully, because selecting one or two evidence channels from noisy EHR data would generally be insufficient unless those channels were strongly justified for the intended task and population. Specialist roles would therefore need to be defined around clinically meaningful and empirically validated evidence channels, with explicit attention to signal quality, missingness, redundancy, and complementarity. It is also possible that EHR-scale data may provide opportunities not captured by this study’s tabular benchmarks. LLMs may benefit from richer semantic signals in clinical notes, narrative descriptions, temporal context, and clinician-authored summaries, rather than relying only on isolated numeric variables. In such settings, role decomposition could potentially support more meaningful specialization, for example by separating structured laboratory evidence from narrative symptom descriptions or longitudinal clinical context. However, this potential should not be assumed from the present results. The appropriate translation of this study is therefore at the level of design principle: role structure and architectural configuration can materially reshape operating characteristics, but EHR-scale systems would require dedicated validation before clinical utility could be established.

### 4.4. Relationship to Prior Work

The observed behavioral differences align with modular design principles from ensemble learning and mixture-of-experts frameworks. Like ensemble systems, the GD and FS protocols combine multiple intermediate opinions before producing a final decision; however, unlike conventional ensembles, the agents in this study share the same underlying model weights and differ only in prompt role and information access [8]. The comparison therefore isolates role-conditioned evidence routing rather than diversity from separately trained models. Similarly, in sparsely activated mixture-of-experts models, routing determines which submodules contribute to a prediction and can change the effective input–output mapping without any weight updates [18]. The present results extend this idea to prompt-level multi-agent systems: even with fixed model parameters, changing how evidence is partitioned across roles can alter aggregation behavior and shift operating characteristics.

Recent clinical LLM studies in oral-lesion diagnosis similarly show that apparent diagnostic performance can vary with prompt design, task formulation, input modality, and evaluation context [19, 20]. These findings support the broader premise that LLM clinical behavior should be evaluated as a function of workflow and task design rather than treated as a fixed property of the underlying model. Evidence from multi-agent prompting indicates that explicit role decomposition can alter task performance and output behavior relative to single-agent baselines. Multi-agent simplification workflows have been shown to improve performance over single-agent approaches on the same dataset, with gains attributed to staged role partitioning and validation rather than changes to the underlying model [21]. In multi-agent reinforcement learning, architectural asymmetry has likewise been shown to influence outcomes, including the introduction of specialized agents within value-decomposition frameworks in heterogeneous environments [22].

Workflow-level analyses of LLM chaining further emphasize that topology affects controllability, transparency, and output characteristics [6]. However, many prior comparisons confound architectural structure with concurrent changes in prompts, model configuration, or training. In contrast, the primary comparison in the present study isolates internal role structure under fixed weights, deterministic decoding, matched computational budget, and shared adjudication logic, while the ablation analyses separately evaluate sensitivity to specialist feature assignment and model capacity.

### 4.5. Limitations

This study evaluates role-structured prompting on two publicly available benchmark datasets (UCI Cleveland Heart Disease and Pima Indians Diabetes) under binary classification, limiting generalizability to other populations, multiclass tasks, and higher-dimensional EHR settings. Both datasets are relatively small and curated, and may not reflect real-world clinical noise, missingness patterns, temporal drift, missing-data mechanisms, or distributional shift. Consequently, the findings should be interpreted as evidence of architecture-induced operating-characteristic shifts under controlled benchmark conditions, not as evidence of clinical deployment readiness.

The study also reports observed full-dataset benchmark metrics without confidence intervals or repeated resampling, so the precision of estimated performance differences across alternative samples was not quantified.

The agent roles used in this study should not be interpreted as clinical workflow roles or substitutes for physician diagnostic reasoning. They are experimental prompt roles designed to isolate how role decomposition and information routing affect binary disease-status classification behavior under fixed inference conditions. The observed false-positive and false-negative profiles therefore represent operating-characteristic changes in benchmark classification, not validated clinical safety behavior. Before any use in clinical decision support, similar systems would require validation on larger and more representative contemporary EHR-scale cohorts, external or prospective validation, calibration analysis, clinician-in-the-loop evaluation, and explicit safety thresholds matched to the intended clinical use case [16].

Given the public availability of these datasets, prior exposure during pretraining of the underlying models cannot be excluded; however, each protocol comparison used identical base weights across agents, so any such exposure would affect both GD and FS within a given model condition. The primary experiments used an 8B locally deployable model to reflect realistic institutional constraints in privacy-sensitive settings, and the additional 14B ablation tested whether protocol-level effects were specific to the original `llama3.1:8b` configuration. However, the study does not evaluate a broad range of model families, instruction-tuning regimes, or frontier-scale models, so the results support model-capacity interaction but do not establish scale-invariant behavior across the wider LLM ecosystem. All experiments used deterministic decoding with fixed aggregation logic, and no calibration analysis, uncertainty estimation, probabilistic decoding, or external validation on independent cohorts was performed. This limits assessment of whether model confidence would be reliable for risk-aware clinical decision support, where calibrated uncertainty is important for triage thresholds, clinician review, and escalation of uncertain cases.

The feature-assignment ablations also show that specialist evidence routing is a limitation as well as a design lever within the FS architecture. FS should therefore be treated as a configurable architecture requiring explicit feature-assignment justification rather than as a feature-agnostic protocol. Poorly chosen, weak, or noisy evidence channels may shift sensitivity–specificity balance in undesirable directions, reinforcing that multi-agent role structure requires explicit validation rather than an assumption of safety or robustness.

## 5. Conclusions and Future Work

This study presented a controlled comparison of two deterministic multi-agent architectures for binary clinical classification across the UCI Cleveland Heart Disease and Pima Indians Diabetes datasets. In the primary comparison, the Generic Deliberative (GD) and Feature-Specialist (FS) protocols differed only in internal role decomposition and information routing under fixed model and inference settings. Across both datasets, altering role structure produced material shifts in operating characteristics within each task. On Cleveland, FS increased overall accuracy and macro-F1 relative to GD while shifting the operating point toward greater specificity and reduced sensitivity. On Pima, role decomposition again altered class-wise behavior, but in the opposite direction, producing a pronounced increase in sensitivity and reduction in specificity.

Feature-assignment and model-scale ablations refine this interpretation. FS behavior was not invariant to the selected specialist features: alternative feature pairs changed both performance and sensitivity–specificity balance, with weak or noisy assignments degrading performance and sometimes producing extreme class bias. The model-scale ablation further showed that GD–FS differences interact with base-model capacity, with the larger 14B configuration producing different operating characteristics from the original 8B configuration. These findings indicate that internal role decomposition can function as a configurable architectural bias rather than a universally beneficial or feature-agnostic design. Its effects are jointly shaped by role structure, specialist feature assignment, adjudication logic, and base-model capacity.

Future work should validate these effects across additional cohorts and structured clinical benchmarks with external testing, extend the comparison to multiclass and higher-dimensional EHR tasks, and examine calibration and probabilistic decoding behavior. Broader model-scale experiments across multiple model families are needed to determine whether architecture-induced operating-characteristic shifts persist across different instruction-tuning regimes and capacity levels. Additional feature-routing studies should evaluate systematic criteria for specialist assignment, including domain expertise, statistical feature importance, redundancy, and complementarity. Agent-count and role-ablation studies may further clarify how architectural granularity influences operating characteristics. Ultimately, structured LLM decision systems should be evaluated not only by aggregate accuracy, but also by how role decomposition, evidence routing, and model capacity redistribute clinically meaningful errors.

## Data Availability

All data used in this study are publicly available from the UCI Machine Learning Repository (Heart Disease Cleveland dataset) and the Pima Indians Diabetes dataset. No new human data were generated. Code and derived outputs necessary to reproduce the results are available at https://github.com/CallumGA/single-vs-multi-agent-llm-diagnostics.git

https://archive.ics.uci.edu/dataset/45/heart+disease

https://github.com/CallumGA/llm-role-decomposition-clinical-classification

https://www.kaggle.com/datasets/uciml/pima-indians-diabetes-database

## Appendices

## Appendix A. Full Prompt Specifications

This appendix provides the exact zero-shot prompt specifications, patient record rendering format, and fallback behavior used for all experimental conditions (single-agent, generic multi-agent, and specialist multi-agent). Prompts are reproduced verbatim to ensure reproducibility. The JSON key `diagnosis` is retained because it was the implementation field name used in the original prompt templates; throughout the manuscript, these outputs are interpreted as benchmark disease-status classification labels rather than clinical diagnoses.

### Appendix A.1. Patient Record Serialization

All models received patient records in a structured text format. The serialization template differed by dataset but preserved a consistent human-readable layout across experiments.

#### UCI Cleveland Heart Disease Dataset

~~~
Patient information:
- Age: {age_years}
- Sex: {sex}
- Chest pain type: {chest_pain_type}
- Resting blood pressure:
     {resting_blood_pressure_mmHg} mmHg
- Serum cholesterol: {serum_cholesterol_mg_dL} mg/dL
- Fasting blood sugar > 120 mg/dL:
     {fasting_blood_sugar_over_120_mg_dL}
- Resting ECG result: {resting_ecg_result}
- Maximum heart rate achieved: {max_heart_rate_bpm}
     bpm
- Exercise-induced angina: {exercise_induced_angina}
- ST depression induced by exercise:
     {st_depression_exercise_relative_to_rest}
- ST segment slope during peak exercise:
     {st_segment_slope_peak_exercise}
- Number of major vessels (fluoroscopy):
     {num_major_vessels_fluoroscopy}
- Thalassemia test result: {thalassemia_test_result}
~~~

#### Pima Indians Diabetes Dataset

~~~
Patient information:
- Age: {age_years}
- Pregnancies: {pregnancies}
- Plasma glucose concentration (2-hour OGTT):
     {plasma_glucose_mg_dL} mg/dL
- Diastolic blood pressure:
     {diastolic_blood_pressure_mmHg} mmHg
- Triceps skinfold thickness:
     {triceps_skinfold_thickness_mm} mm
- 2-hour serum insulin: {serum_insulin_uU_mL} uU/mL
- Body mass index (BMI): {body_mass_index} kg/m^2
- Diabetes pedigree function:
     {diabetes_pedigree_function}
~~~

### Appendix A.2. Single-Agent Baseline Prompt

~~~
You are a clinician performing a binary diagnostic
     classification task.
You will receive the full patient record.
Your job:
Return the final diagnosis:
- “disease_present” or “no_disease”
Rules (do not mention these in output):
- Consider all relevant factors in the patient record.
- If evidence clearly supports disease, return
    “disease_present”.
- Otherwise return “no_disease”.
- If genuinely uncertain, choose “no_disease”.
Output MUST be a single JSON object and nothing else.
{ “diagnosis”: “disease_present” OR “no_disease” }
~~~

### Appendix A.3. Generic Deliberative (GD) Protocol

#### Appendix A.3.1. Doctor Opinion Prompt

~~~
You are a clinician reviewing the FULL patient record.
Task:
Provide an overall opinion about whether the FULL
      RECORD supports:
- supports_disease
- supports_no_disease
- unclear
Guidelines (do not mention these in output):
- Use “supports_disease” only if overall evidence
      clearly supports disease.
- Use “supports_no_disease” only if overall evidence
      clearly supports no disease.
- Otherwise use “unclear”.
- “strength” reflects how strong the overall evidence
      is (high > medium > low).
- “note” must be ONE short sentence citing 1 to 2 key
      factors from the record (no long reasoning).
Output MUST be a single JSON object and nothing else:
{
    “scope”: “overall_record”,
    “signal”: “supports_disease” OR
       “supports_no_disease” OR “unclear”,
    “strength”: “low” OR “medium” OR “high”,
    “note”: “one short sentence”
}
~~~

#### Appendix A.3.2. Adjudicator Prompt

~~~
You are an adjudicator for a binary clinical
      classification task.
You will receive:
1) The full patient record
2) Two doctor opinions about the full record (signal
      + strength)
Your job:
Return the final diagnosis:
- “disease_present” or “no_disease”
Rules (do not mention these in output):
- Treat each doctor opinion as evidence about the
      overall record; it may be low quality or wrong.
- Weigh evidence by “strength” (high > medium > low).
- If both doctors strongly support the same
      direction, follow that direction.
- If evidence conflicts or is weak/unclear, use the
      full patient record to decide.
- If still genuinely uncertain after considering the
      full record, choose “no_disease” as a tie-break.
- Do not automatically prefer Doctor 1 or Doctor 2.
Output MUST be a single JSON object and nothing else.
     { “diagnosis”: “disease_present” OR “no_disease” }
~~~

### Appendix A.4. Feature-Specialist (FS) Protocol Appendix

#### A.4.1. Feature Specialist Prompt

~~~
You are a clinical feature specialist. You ONLY
evaluate the feature: “{feature_name}”.
Task:
Given the patient record, decide whether THIS FEATURE
      ALONE:
- supports_disease
- supports_no_disease
- unclear
Guidelines (do not mention these in output):
- Use “supports_disease” only if this feature value
      is a meaningful risk indicator by itself.
- Use “supports_no_disease” only if this feature
      value is meaningfully reassuring by itself.
- Otherwise use “unclear”.
- “strength” reflects how strongly this single
      feature points in that direction (not overall
      diagnosis):
- high: strong / clear signal
- medium: moderate signal
 - low: weak, borderline, missing, or noisy
- “note” must be ONE short sentence that explicitly
      cites the patients “{feature_name}” value and why
      it points that way.
- Do NOT use any other features in your reasoning.
Output MUST be a single JSON object and nothing else,
      with EXACTLY these keys:
{
    “feature”: “{feature_name}”,
    “signal”: “supports_disease” OR
        “supports_no_disease” OR “unclear”,
    “strength”: “low” OR “medium” OR “high”,
    “note”: “one short sentence referencing the
        patient’s {feature_name} value”
}
~~~

#### Appendix A.4.2. Specialist Adjudicator Prompt

~~~
You are an adjudicator for a binary clinical
      classification task.
You will receive:
1) The full patient record
2) Two specialist opinions (each about ONE feature
      only)
Your job:
Return the final diagnosis:
- “disease_present” or “no_disease”
Rules (do not mention these in output):
- Treat each specialist opinion as evidence about a
      single feature; it may be low quality or wrong.
- Weigh evidence by “strength” (high > medium > low).
- If both specialists strongly support the same
      direction, follow that direction.
- If evidence conflicts or is weak/unclear, use the
      full patient record to decide.
- If still genuinely uncertain after considering the
      full patient record, choose “no_disease” as a
      tie-break.
Output MUST be a single JSON object and nothing else.
{ “diagnosis”: “disease_present” OR “no_disease” }
~~~

## Appendix B. Full Feature Mapping

This study uses two publicly available datasets from the UCI Machine Learning Repository: the UCI Heart Disease dataset (Cleveland subset) [12] and the Pima Indians Diabetes dataset [13]. Feature definitions and categorical encodings follow the official dataset documentation distributed with each download.

### UCI Heart Disease (Processed Cleveland Subset)

Table B.1 lists the full feature schema and encodings used in this study.

### Pima Indians Diabetes Dataset

Table B.2 lists the full feature schema and encodings used for the Pima dataset.

## Funding

No external funding was received for this work.

## Conflicts of Interest

The author declares no competing interests.

## Ethics Statement

This study uses publicly available, fully de-identified datasets: the UCI Heart Disease (Cleveland subset) dataset and the Pima Indians Diabetes dataset. No new data were collected and no human participants were recruited. As the analysis relied exclusively on secondary use of anonymized public data, institutional review board approval was not required.

## Data and Code Availability

The datasets used in this study are publicly available: UCI Heart Disease dataset (Cleveland subset): https://archive.ics.uci.edu/dataset/45/heart+disease

Pima Indians Diabetes dataset (Kaggle version): https://www.kaggle.com/datasets/uciml/pimaindians-diabetes-database

All source code, prompt templates, and evaluation scripts are publicly available at: https://github.com/CallumGA/llm-multi-agent-role-decomposition.

**Table B.1:**
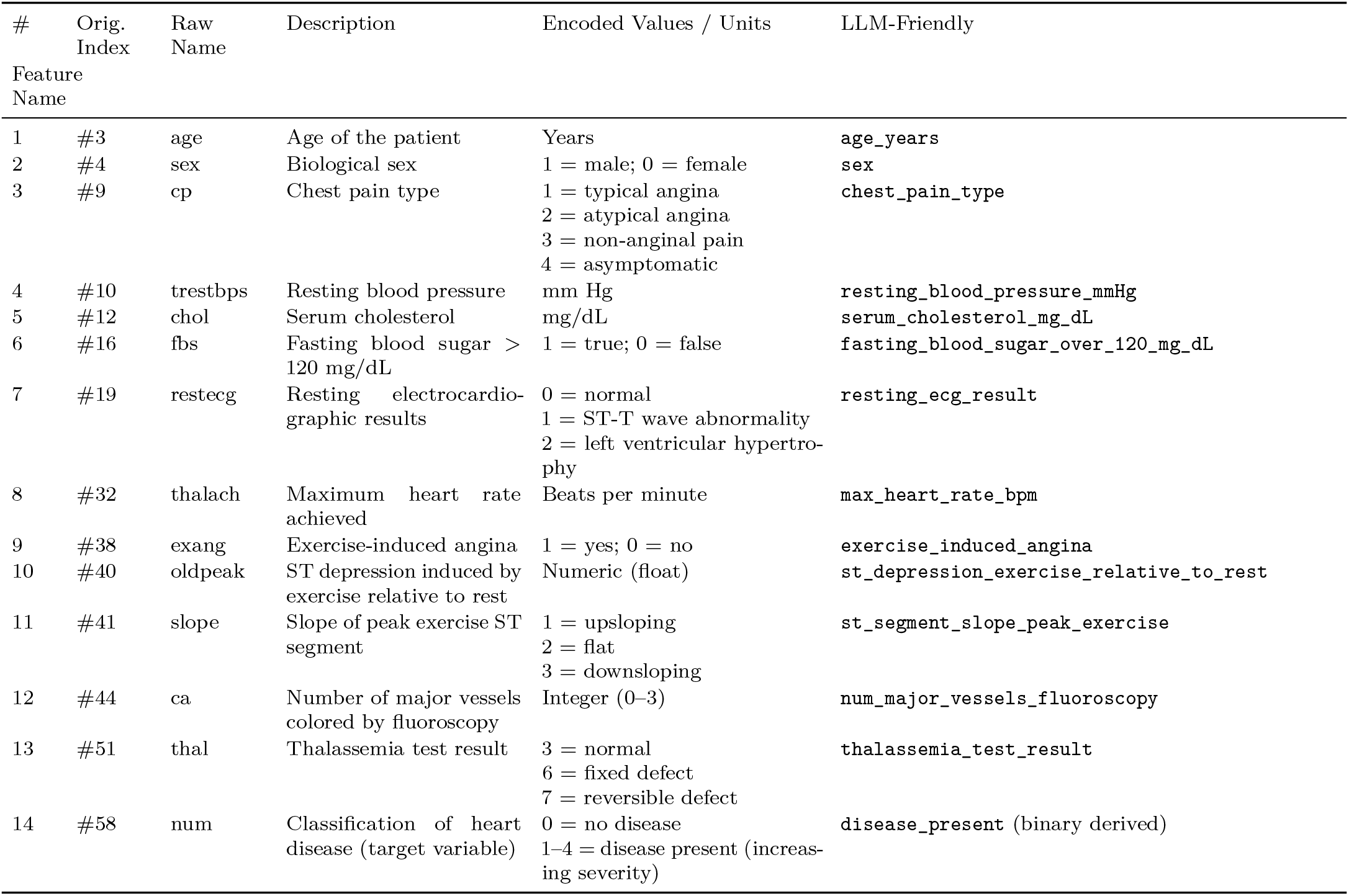
Full feature schema and categorical encodings for the UCI Heart Disease (Processed Cleveland subset).

**Table B.2:**
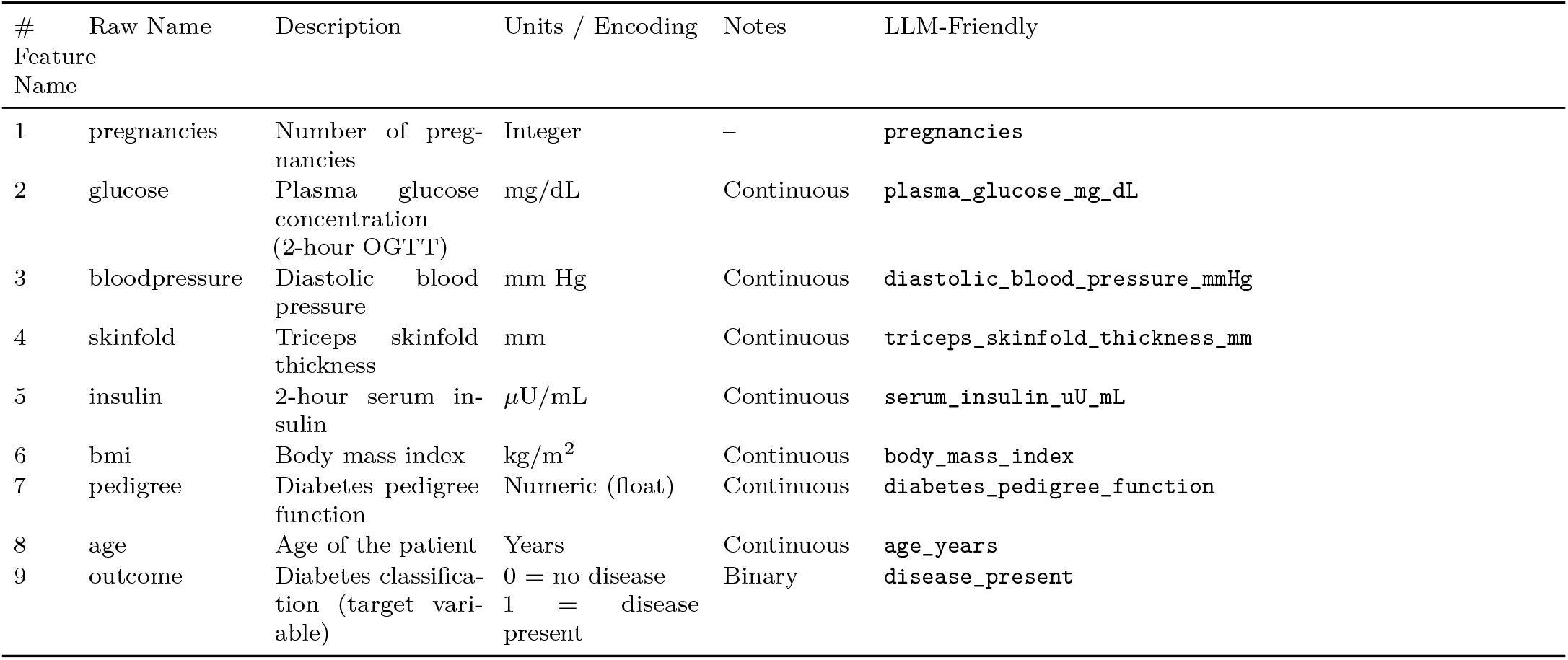
Full feature schema and encodings for the Pima Indians Diabetes dataset.

## Notes

### Competing Interest Statement

The authors have declared no competing interest.

### Funding Statement

This study did not receive any external funding. The authors did not receive payment or services from any third party for any aspect of the submitted work, including study design, data analysis, manuscript preparation, or publication.

### Author Declarations

The study used only publicly available, fully de-identified human data from the UCI Machine Learning Repository. Specifically, the Cleveland Heart Disease dataset was obtained from: UCI Machine Learning Repository Heart Disease Dataset (Cleveland) https://archive.ics.uci.edu/dataset/45/heart+disease

### Summary of Updates

The manuscript was substantially revised in response to reviewer comments. The main changes include new feature-assignment sensitivity analyses for the Feature-Specialist protocol across both datasets, a model-scale sensitivity analysis using a 14B model, and expanded discussion of clinical interpretation, sensitivity-specificity trade-offs, and translation to real-world EHR settings. The manuscript now frames the findings as controlled benchmark disease-status classification results rather than evidence of clinical deployment readiness. Agent-role terminology, preprocessing choices, fallback reporting, and disease-status classification language were also clarified. The Abstract, Discussion, and Conclusions were softened to emphasize that role decomposition should be interpreted as a feature-conditioned architectural effect rather than a universally beneficial or model-independent mechanism.

## References

[1] D. Fernandes Prabhu, V. Gurupur, A. Stone, E. Trader, Integrating artificial intelligence, electronic health records, and wearables for predictive, patient-centered decision support in healthcare, Healthcare 13 (21) (2025) 2753. doi:10.3390/healthcare13212753.

[2] A. Sarker, R. Zhang, Y. Wang, Y. Xiao, S. Das, D. Schutte, D. Oniani, Q. Xie, H. Xu, Natural language processing for digital health in the era of large language models, IMIA Yearb. Med. Inform. (2024). doi:10.1055/s-0044-1800750.

[3] A. Bornet, P. Khlebnikov, F. Meer, Q. Haas, A. Yazdani, B. Zhang, P. Amini, D. Teodoro, Analysis of eligibility criteria clusters based on large language models for clinical trial design, J. Am. Med. Inform. Assoc. 32 (3) (2025) 447–458. doi:10.1093/jamia/ocae311.

[4] J. C. L. Ong, B. J. J. Seng, J. Z. F. Law, L. L. Low, A. L. H. Kwa, K. M. Giacomini, D. S. W. Ting, Artificial intelligence, chatgpt, and other large language models for social determinants of health: current state and future directions, Cell Rep. Med. 5 (2024) 101356. doi:10.1016/j.xcrm.2023.101356.

[5] A. Neveditsin, et al., A comprehensive review of language models in medicine, PLOS Digit. Health 4 (5) (2025) e0000800. doi:10.1371/journal.pdig.0000800.

[6] M. Grunde-McLaughlin, M. S. Lam, R. Krishna, D. S. Weld, J. Heer, Designing LLM chains by adapting techniques from crowdsourcing workflows, ACM Trans. Comput.-Hum. Interact. 32 (3) (2025) 27. doi:10.1145/3716134.

[7] G. Li, H. A. A. K. Hammoud, H. Itani, D. Khizbullin, B. Ghanem, CAMEL: Communicative agents for “mind” exploration of large language model society, arXiv preprint arXiv:2303.17760 (2023). URL https://arxiv.org/abs/2303.17760

[8] R. Polikar, Ensemble based systems in decision making, IEEE Circuits Syst. Mag. 6 (3) (2006) 21–45. doi:10.1109/MCAS.2006.1688199.

[9] V. Harish, et al., Teaching old tools new tricks— preparing emergency medicine for the impact of machine learning-based risk prediction models, CJEM 25 (5) (2023) 365–369. doi:10.1007/s43678-023-00480-8.

[10] Y. Chen, B. Wen, F. Zulkernine, A Multi-agent summarization and auto-evaluation framework for medical text: Development and evaluation study, JMIR AI 4 (2025) e75932. doi:10.2196/75932.

[11] R. Perera, A. Basnayake, M. Wickramasinghe, Autoscaling LLM-based multi-agent systems through dynamic integration of agents, Front. Artif. Intell. (2025). doi:10.3389/frai.2025.1638227.

[12] D. Dua, C. Graff, Heart disease data set [dataset], UCI Machine Learning Repository, accessed 2026; original data donated in 1988 (2017). URL https://archive.ics.uci.edu/dataset/45/heart+disease

[13] UCI Machine Learning Repository, Pima indians diabetes database [dataset], Kaggle, originally from the UCI Machine Learning Repository; accessed 2026 (2016). URL https://www.kaggle.com/datasets/uciml/pima-indians-diabetes-database

[14] R. R. Sarra, A. M. Dinar, M. A. Mohammed, Enhanced heart disease prediction based on machine learning and *χ*2 statistical optimal feature selection model, Designs 6 (5) (2022) 87. doi:10.3390/designs6050087.

[15] H. Zhou, Y. Xin, S. Li, A diabetes prediction model based on boruta feature selection and ensemble learning, BMC Bioinformatics 24 (1) (2023) 224. doi: 10.1186/s12859-023-05300-5.

[16] L. Wynants, B. Van Calster, G. S. Collins, R. D. Riley, G. Heinze, E. Schuit, M. M. J. Bonten, J. A. A. Damen, T. P. A. Debray, M. De Vos, et al., Prediction models for diagnosis and prognosis of covid-19: systematic review and critical appraisal, BMJ 369 (2020) m1328. doi:10.1136/bmj.m1328.

[17] E. Hossain, R. M. Rana, N. Higgins, J. Soar, P. D. Barua, A. R. Pisani, K. Turner, Natural language processing in electronic health records in relation to healthcare decision-making: A systematic review, Computers in Biology and Medicine 155 (2023) 106649. doi:10.1016/j.compbiomed.2023.106649.

[18] W. Fedus, B. Zoph, N. Shazeer, Switch transformers: Scaling to trillion parameter models with simple and efficient sparsity, J. Mach. Learn. Res. 23 (120) (2022) 1–39. URL https://jmlr.org/papers/v23/21-0998.html

[19] F. E. A. Hassanein, M. Alkabazi, M. Tassoker, Y. Ahmed, S. Alsaeed, A. Abou-Bakr, Multimodal large language models for oral lesion diagnosis: a systematic review of diagnostic performance and clinical utility, Front. Oral Health 7 (2026) 1748450. doi:10.3389/froh.2026.1748450. URL https://www.frontiersin.org/journals/oral-health/articles/10.3389/froh.2026.1748450/full

[20] F. E. A. Hassanein, S. S. Ibrahim, S. Tomo, A. Alsahhaf, A. Abou-Bakr, Prompt engineering shapes diagnostic accuracy and explanation quality of LLM in oral lesion diagnosis: a prospective, expert-blinded benchmark study, Odontology (2026). doi:10.1007/s10266-026-01380-w. URL https://link.springer.com/article/10.1007/s10266-026-01380-w

[21] P. Zunjare, M. S. Hsiao, A hybrid multi-agent prompting approach for simplifying complex sentences, in: 2025 IEEE Smart World Congress (SWC), IEEE, 2025. doi:10.1109/SWC65939.2025.00070.

[22] Q. Kang, F. Wang, Z. Liu, Z. Chen, Special agents policy gradient in value decomposition-based approach, in: 2023 IEEE 12th Data Driven Control and Learning Systems Conf. (DDCLS), IEEE, 2023. doi:10.1109/DDCLS58216.2023.10165847.

